# When Words Sound Alike: Decoding Homophone Processing in Chinese Primary Progressive Aphasia

**DOI:** 10.64898/2026.10.16.26364895

**Authors:** Boon Lead Tee, Ta-Fu Chen, Lorinda Kwan-Chen Li-Ying, YuChen Chuang, Raymond Y. Lo, Joshua Tsoh, Andrew Lung-Tat Chan, Adrian Wong, Vincent Mok, Tin Yee Connie Yan, Chien Jung Lu, Yu Sun, YiChen Lee, Isabel Elaine Allen, FangDa Leng, Yu-Wen Cheng, Chin-Hsien Lin, Cheng-Hsuan Li, Yu Chen, GuoYu Zhou, Adeline Ng, Jet Vonk, Maria Luisa Gorno-Tempini

## Abstract

Chinese is one of the living languages with the highest homophone densities, making homophone processing a critical component of language comprehension and orthographic decoding, as well as an ideal model for examining its cognitive mechanisms. This study assessed mono- and bi-character homophone selection function in 129 Mandarin- and Cantonese-speaking participants, including 66 cognitively healthy individuals, 17 with semantic variant (sv) primary progressive aphasia (PPA), 23 with the nonfluent/agrammatic variant (nfv) PPA, and 23 with the logopenic variant (lv) PPA. All PPA variants showed impairment in homophone selection, with most significant deficits in svPPA and lvPPA. ROC analyses demonstrated its strong diagnostic utility for distinguishing PPA from controls. Voxel-based morphometry revealed that homophone performance correlated with gray-matter volume in left anterior and inferior temporal regions, with variant-specific patterns observed. These findings described the potential role of homophone selection in diagnosing PPA syndrome in Chinese speakers, illuminate cross-linguistic influences on language impairment in PPA, and provide novel insights into the cognitive processing of homophones.

## 1. Introduction

Primary Progressive Aphasia (PPA) is a clinical syndrome characterized by insidious onset progressive deterioration of speech and language functions, typically associated with focal degeneration in left-hemispheric speech and language networks. PPA is commonly categorized into three canonical variants that differ not only in their cognitive-linguistic profiles but also in their characteristic neuroanatomical and pathological distributions. Semantic variant PPA (svPPA), characterized by profound degradation of conceptual knowledge, is associated with anterior and inferior temporal atrophy, typically more pronounced in the left hemisphere, and is most commonly linked to TDP-43 type C pathology. Nonfluent/agrammatic variant PPA (nfvPPA), marked by agrammatism and/or apraxia of speech, predominantly affects the left inferior frontal, insular, premotor, and supplementary motor regions and is frequently associated with tauopathy. Logopenic variant PPA (lvPPA), characterized by impairment in lexical retrieval, phonological and orthographic processing, and auditory verbal short term memory, is associated with left posterior temporal and inferior parietal degeneration and is most commonly underpinned by Alzheimer’s disease pathology.(Gorno-Tempini et al., 2011) Nevertheless, clinical manifestation of PPA has been shaped largely by studies of Germanic and Romance language speakers, particularly English.(García et al., 2023) Given that more than 7,000 languages are spoken worldwide, it is essential to exercise caution when generalizing these clinical findings to linguistic systems that differ substantially from Indo-European languages.(Dryer & Haspelmath, 2013; García et al., 2023) Incorporating typologically informed linguistic features into assessment is therefore critical for improving diagnostic precision in non-Indo-European language speakers with PPA.

Homophone processing exemplifies one such feature. Homophones, defined as words that share identical pronunciation but differ in meaning and orthography, represent a salient aspect of lexical-semantic organization that intersects with orthographic and phonological processing. Typical examples of homophones in English include words such as eight-ate and write-right-wright-rite. In languages with high homophone density (i.e., the number of distinct lexical items sharing the same pronunciation), such as Chinese languages, cognitive processing of homophones plays a particularly prominent role in speech and language functions. While English words typically have only two to three homophonic counterparts, Mandarin Chinese words exhibit a markedly higher degree of phonological overlap, averaging 8 to 11 homophones per monosyllabic word, with some syllables associated with over 50 distinct homophonic forms.(Tan & Perfetti, 1998; Wang et al., 2012; Yao & Sharma, 2017) For instance, for the Mandarin monosyllable /Yì/ (with the diacritic indicating Tone 4), we identified 82 distinct characters sharing the same pronunciation but differing in semantic meaning and orthography (Supplement Table 1). The estimated homophone density in Mandarin also varies considerably depending on the size of the lexical corpus and the unit of measurement, specifically whether homophony is calculated at the level of monosyllabic words (e.g., sea - see in English; 中 “middle” /zhōng/- 忠 “loyal” /zhōng/ in Mandarin; 游 “swim” /jau4/- 油 “oil” /jau4/ in Cantonese) or bi-/multisyllabic compound words (e.g., foreword - forward in English; 店員 “shop asssistant” /diànyuán/- 電源 “power supply” /diànyuán/ in Mandarin; 全民 “entire population” /cyun4man4/-傳聞 “rumor” /cyun4man4/ in Cantonese). Early dictionary-based analyses reported that approximately 11-15% of Mandarin words are homophonous, compared with roughly 3.2% of English words.(Neergaard et al., 2022; Wen, 1980) The more recent corpus-based studies suggest substantially higher rates, with 20-40% of monosyllabic words sharing their pronunciation with other characters within the lexicon.(Sun et al., 2018; Wang et al., 2012; Yao & Sharma, 2017) In spoken Chinese, high homophone density generates substantial lexical ambiguity, making context essential for identifying the intended meaning. Resolving this ambiguity requires the sentence context to be integrated with the distinct orthographic and semantic representations of competing phonologically identical words. (Chen et al., 2009; Li & Yip, 1998; Yao & Sharma, 2017)

Among cognitively healthy individuals, homophone density has been demonstrated to influence lexical-semantic decision-making in both auditory and visual domains. (Bao et al., 2024; W. F. Chen et al., 2016; Wang et al., 2012) Eye-tracking studies further indicate that native Mandarin and Cantonese listeners use sentence context to anticipate and preferentially activate the intended homophones.(Yip & Zhai, 2018) Electrophysiological findings further demonstrate that homophone density and orthographic consistency modulate neural responses during Chinese spoken-word recognition.(W. F. Chen et al., 2016) Complementary neuroimaging research on lexical ambiguity, including homonymy and polysemy, implicates frontal-temporal systems in resolving lexical-semantic competition.(Bedny et al., 2008; Hoffman & Tamm, 2020; Rodd et al., 2012) Collectively, these neuroimaging findings suggest that contextual homophone selection depends on the coordinated engagement of orthographic, semantic, and cognitive-control systems rather than a single homophone-specific region.

While homophone processing is a central linguistic component in numerous East and Southeast Asian languages, including Mandarin, Cantonese, Vietnamese, Thai, Japanese, and Korean, there has been limited research examining this phenomenon in individuals with neurological disorders.(Biedermann et al., 2002) Investigating this linguistic feature in homophone-rich languages offers a valuable opportunity to elucidate how specific linguistic structures are differentially affected in PPA across languages and may improve diagnostic precision for non-Indo-European language speakers. Chinese languages are uniquely well suited for investigating homophone function, owing to their exceptionally high degree of homophonous lexicons. The frequent need to disambiguate meaning through contextual and orthographic cues renders Chinese an especially informative model for elucidating the cognitive and neural mechanisms underlying homophone processing.

In this study, we examined homophone selection function in Chinese speakers with PPA. Participants are tasked to select the contextually appropriate written word from phonologically identical alternatives that differed in orthography and meaning. As phonological information alone cannot distinguish the candidates, accurate selection requires access to their distinct orthographic and semantic representations and integration of this lexical information with the disambiguating sentence context. We pursued three objectives in this study. First, we characterized homophone selection performance across PPA variants. We hypothesized that performance would be impaired in svPPA because of semantic knowledge degradation, in lvPPA because of orthographic processing impairment, and in nfvPPA because of sentence-level comprehension difficulties. We also examined these differences between mono- and bi-character homophones, which place distinct demands on lexical competition and morphological composition, and between Mandarin and Cantonese. The latter comparison was motivated by the greater divergence between colloquial speech and formal written Chinese in Cantonese, a diglossic language. For example, “He does not like eating fish” is formally written as 他不喜歡吃魚, closely corresponding to its spoken Mandarin form, whereas colloquial Cantonese typically expresses the same meaning verbally as 佢 唔鍾意食魚, using different forms for the pronoun, negation, preference, and eating. This spoken-written divergence may place additional demands on integrating phonological, orthographic, and semantic representations in Cantonese. Second, to evaluate its potential clinical relevance, we examined whether homophone selection accuracy distinguished individuals with PPA from cognitively healthy controls and contributed discriminative information beyond demographic characteristics. We further compared its performance with conventional neuropsycholinguistic measures, hypothesizing that homophone selection would afford strong discrimination without necessarily serving as a superior or standalone diagnostic instrument. Third, we investigated the cognitive and neural mechanisms supporting homophone processing. We hypothesized that greater homophone selection accuracy would be associated with preserved gray-matter volume in regions supporting semantic knowledge, orthographic representation, and sentence processing. Guided by the characteristic anatomical profiles of the PPA variants and the cognitive processes we hypothesized that impairment would be associated with degeneration of anterior and inferior temporal regions supporting semantic representation in svPPA; posterior temporal and ventral occipitotemporal regions supporting lexical retrieval and orthographic representation in lvPPA; and inferior frontal regions supporting sentence processing and cognitive control in nfvPPA. Together, this work offers novel neurocognitive insights into homophone processing and enriches our cross-linguistic understanding on how variation in linguistic structures influence PPA symptomalogy.

## Material and Methods

### Participants

This study included a total of 129 native Chinese speakers who were enrolled into the international Chinese Language Assessment in Primary Progressive Aphasia (CLAP) project, in which recruitment was conducted across seven collaborating sites in Hong Kong, Taiwan, and the United States.(Tee, DeLeon, et al., 2022; Tee et al., 2024; Tee, Kwan-Chen, et al., 2022) The cohort comprised 66 cognitively healthy individuals and 63 patients who fulfilled current consensus diagnostic criteria for primary progressive aphasia (PPA), including 17 with svPPA, 23 with nfvPPA, and 23 with lvPPA. All included participants completed the homophone selection assessment as part of the CLAP neuropsycholinguistic assessment battery.(Gorno-Tempini et al., 2011) To ensure adequate familiarity with Chinese languages, participants were required to have completed at least six years of schooling conducted in Chinese. Those with fewer than six years of formal schooling were eligible only if their education had been conducted entirely in Chinese. Participants were excluded if they had other neurodegenerative diseases, a history of major brain trauma or neurosurgery, oropharyngeal conditions affecting articulation, significant visual or hearing impairment, or a Mini-Mental State Examination (MMSE)(Folstein et al., 1975) lower than 10. The study protocol received approval from the institutional review boards at all participating institutions.

### Neuropsycholinguistic assessment

We collected MMSE, CLAP neuropsycholinguistic assessment, and brain MRI imaging data from the enrolled participants. As detailed in previous studies,(Tee et al., 2024; Tee, Kwan-Chen, et al., 2022) the CLAP neuropsycholinguistic assessment battery, comprises numerous speech and language tests, including confrontation naming, single-word comprehension, semantic association, motor speech, and sentence repetition tests. In the CLAP confrontation naming test, participants were tasked with naming 48 pictured items with high concreteness but varying word frequency. In the single-word comprehension test, participants were provided the auditory names of 15 objects or animals and were asked to select the correct image from six options within the same semantic category. The CLAP semantic association tests comprised 30 triads of words or pictures, with the target stimulus presented above two choices. Participants were then asked to identify the option most semantically related to the target. To assess motor speech skills, we tasked participants to repeat for five times three distinct sets of phrases, each consisted of four- or six-characters.

These phrases were systematically designed to assess specific articulatory features: (1) Place of articulation: phrases that varied in articulatory placement while holding tone and manner constant; (2) Manner of articulation: phrases that differed in how sounds are produced, with consistent articulatory placement and tone across syllables; (3) Tone of articulation: phrases that presented tonal contrasts across otherwise syllabically similar words. In the repetition task, participants were tasked to repeat both semantically coherent and semantically anomalous phrases and sentences, ranging from three to eleven Chinese words in length. Orthographic ability was assessed using a 34-item abbreviated version of the CLAP orthographic dictation test. Each target character was presented auditorily within a two-character word to disambiguate its homophones, and accuracy was calculated as the percentage of correctly written characters.(Tee, Kwan-Chen, et al., 2022) The syntax comprehension test comprised 30 Chinese sentences describing interactions between two characters, with syntactic complexity systematically varied across items. Each sentence was accompanied by two or four illustrations, and participants selected the picture that best matched its meaning; 23 items contained syntactic foils and seven contained semantic foils, with accuracy expressed as the percentage of correct responses. Assessments were conducted in each participant’s dominant language (Cantonese or Mandarin), as determined by language proficiency and usage scores using language history questionnaire 3.0.(Li et al., 2020) All assessments were administered by board-certified neurologists, neuropsychologists, speech-language pathologists, or supervised research staff, all of whom are native Mandarin or Cantonese speakers with over 12 years of formal education in the Chinese languages.

### Homophone selection

The homophone selection test was designed to assess participants’ ability to select semantically and orthographically appropriate homophone items from sets of phonologically and tonologically identical options. The task employs a sentence completion paradigm, in which participants fill in a missing mono-character (e.g., 騎) or bi-character (e.g., 騎士) compound word in a short sentence by choosing the correct homophone from a set of visually presented options (test items and stimuli are provided in Supplementary Table 2). For each trial, the sentence context and all candidate homophones were visually presented simultaneously. For example, the Mandarin sentence 汽車撞倒了___士 (“The car knocked down the ___rider”) required participants to select 騎 /qí/ (“ride”) from the homophonic alternatives旗,(“flag”), 奇 (“strange”), and 齊 (“together/even”), all pronounced /qí/ but differing in orthography and meaning. The selected character, representing a single morpheme, combined with 士, to form the compound word 騎士 /qíshì/, meaning “motorcyclist.” An approximate English analogue would be selecting SAIL rather than SALE to complete “The rough water capsized the ___BOAT.” In the bisyllabic condition, participants are tasked to select an two-character bisyllabic bimorphemic compound word form, with alternatives sharing its complete bisyllabic pronunciation. For example, the Mandarin sentence ___說: 「歡迎光臨」 (“The ___ said, ‘Welcome’”) required selection of 店員 (“shop assistant”) rather than the homophonous alternative 電源 (“power supply”), both pronounced /diànyuán/. An approximate English analogue would be selecting FOREWORD rather than FORWARD to complete “The distinguished scholar wrote the ___ to the new edition.” In Mandarin, the test comprised 10 mono-character and 10 bi-character homophone trials, while the Cantonese version included 8 mono-character and 8 bi-character trials. All sentence items were constructed to be 5-11 character length and designed to ensure semantic constraint to the target homophone. The inclusion of both mono- and bi-character items allows for the examination of homophone processing at different levels of morphological complexity, capturing performance at the single morpheme/character level as well as at the two morpheme, compound word level.

### Statistical Analysis

We compared demographic characteristics across diagnostic groups using analysis of variance (ANOVA) for continuous measures with Bonferroni correction for multiple comparisons and Pearson’s Chi-squared tests for categorical variables. Group differences in years of education, MMSE scores, and CLAP neuropsycholinguistic performance were evaluated using nonparametric Kruskal–Wallis H tests, as the data did not meet the assumptions of normality and homogeneity of variance based on Shapiro–Wilk tests. Post-hoc pairwise comparisons were conducted with Bonferroni adjustment to control for family-wise error, with the significance level set at α = 0.05 (two-tailed).

Observed total, mono-character, and bi-character homophone selection accuracy across diagnostic groups were evaluated using Kruskal-Wallis tests followed by Bonferroni-adjusted pairwise comparisons. To further account for potential confounding effects of age, education, and testing language (Cantonese vs. Mandarin), group differences in homophone performance (total, monocharacter, and bicharacter word scores) were analyzed using logistic regression models, with the number of correct responses modeled as a binomial outcome relative to the total number of trials. This approach was selected as homophone accuracy scores did not meet the assumptions of normality. Diagnostic group and testing language were specified as fixed factors, with their interaction term included in the model, while age at examination and years of education were entered as covariates. The diagnostic group × testing language interaction examined whether Mandarin–Cantonese differences in homophone selection accuracy varied across diagnostic groups. Given the greater divergence between spoken and written Cantonese and the distinct profiles of impaired and preserved cognitive-linguistic abilities across PPA variants, the processes available to support task performance, and thus the effect of testing language, may differ by diagnostic group. Robust covariance estimates were applied to account for potential heteroscedasticity, and post hoc pairwise comparisons were performed with Šidák adjustment to control for multiple comparisons. All statistical analyses were conducted using IBM SPSS Statistics, version 27.0, with a two-tailed significance threshold set at α = 0.05.

We also conducted receiver operating characteristic (ROC) analyses using the pROC package in R to evaluate the ability of homophone selection accuracy to discriminate PPA patients from cognitively healthy controls. Three complementary analyses were conducted. First, unadjusted ROC curves were generated from the mono-character and bi-character homophone accuracy scores to quantify their discriminative performance. AUCs and 95% confidence intervals were estimated, and optimal raw-score thresholds were determined using Youden’s index. Second, covariate-adjusted ROC models were constructed by deriving predicted probabilities from separate multivariable logistic-regression models incorporating either mono-character or bi-character homophone accuracy together with age at examination, sex, years of education, and testing language (Mandarin or Cantonese). The predicted probabilities were submitted to ROC analysis to estimate the discriminative performance of each multivariable model, and their AUCs were compared using a paired DeLong test restricted to participants with complete data for both models. Third, to determine whether total homophone selection accuracy provided incremental discrimination beyond demographic and language characteristics, a baseline logistic-regression model containing age, sex, education, and testing language was compared with a full model additionally containing total homophone-selection accuracy. The incremental contribution of homophone accuracy was evaluated using a likelihood-ratio test of the nested models and a paired DeLong comparison of their AUCs, with both models estimated in the same complete-case sample. To further contextualize its discriminative performance, the AUC for total homophone selection accuracy was compared with those of the ten neuropsycholinguistic measures in Table 1 using paired nonparametric tests restricted to participants with valid scores on both measures. P values were Holm-adjusted across the ten comparisons, with full results reported in Supplementary Table 4.

**Table 1.**
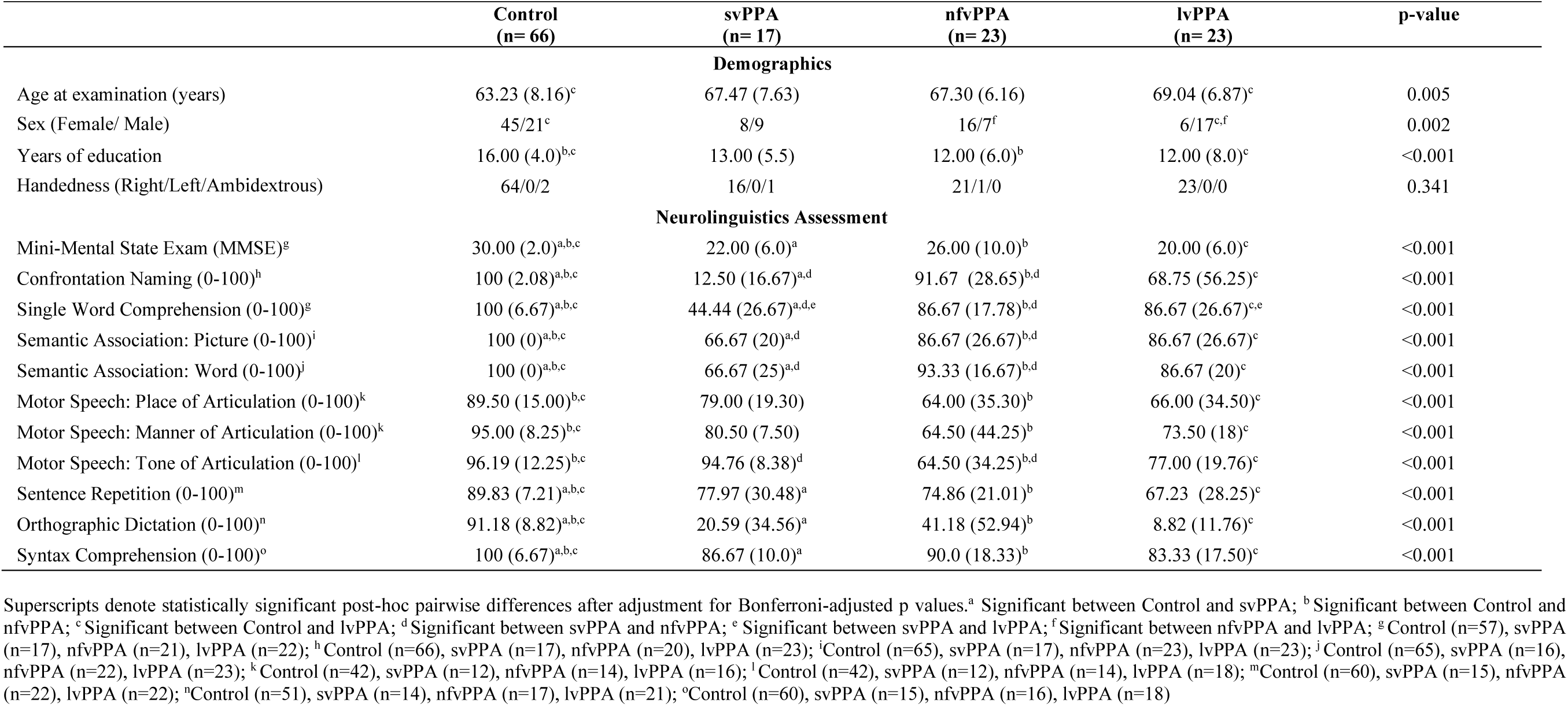
Demographic characteristics and neuropsychological performances of the study participants (n=129).

To further characterize the component processes associated with homophone selection performance, exploratory correlation analyses were conducted within the pooled PPA sample. Homophone-selection accuracy was correlated with percentage scores on ten neuropsycholinguistic measures: confrontation naming, single word comprehension, semantic association: picture, semantic association: word, orthographic dictation, syntax comprehension, sentence repetition, and motor speech measures of place, manner, and tone of articulation. As the percentage scores were bounded and their distributions could not be assumed to be normally distributed, two-tailed Spearman rank-order correlations were used. Missing observations were handled using pairwise deletion; consequently, the sample size for individual correlations ranged from 42 to 58. To control the familywise error rate across the ten correlations, p values were adjusted using the Holm step-down procedure. Statistical significance was evaluated using a two-tailed Holm-adjusted p < 0.05.

As the sample size was determined by participant availability across sites rather than a prespecified recruitment target, we conducted a sensitivity power analysis to estimate the minimum effect detectable for the study’s primary behavioral outcome, total homophone-selection accuracy across the four diagnostic groups. For a four-group omnibus comparison with the present sample (N = 129) at α = 0.05 (two-tailed) and 80% power, the analysis was sensitive to effects of Cohen’s f ≥ 0.30 (η² ≥ 0.08), corresponding to a medium-to-large effect; the observed effect for this outcome substantially exceeded this threshold (η² ≈ 0.50). The secondary and exploratory analysis families incorporated their respective multiple-comparison controls: Bonferroni and Šidák adjustments for post hoc group comparisons, Holm correction for correlation and paired-AUC comparisons, and cluster-level familywise-error correction within each voxelwise model.

### Brain behavioral correlation

To investigate the neural mechanisms underlying homophone performances, we analyzed the neural correlates in participants who completed the CLAP neuropsycholinguistic measures and MRI within a three-month interval. The analysis included 10 participants with svPPA, 12 participants with nfvPPA, 15 patients with lvPPA, and 50 cognitively healthy controls. MRI data were acquired on 3T scanners following the Alzheimer’s Disease Neuroimaging Initiative 3 protocol (ADNI 3; http://adni.loni.usc.edu/adni-3/). The ADNI 3 imaging protocol includes a high-resolution T1-weighted 3D magnetization-prepared rapid gradient-echo (MPRAGE) sequence (200-211 sagittal slices; TR = 2300 ms; TE = 2.98 ms; TI = 900 ms; flip angle = 9°-11°; field of view = 256 mm³; matrix size = 256 × 256; voxel size = 1.0 × 1.0 × 1.0 mm³). T1-weighted images were preprocessed using Computational Anatomy Toolbox (CAT12) in the Statistical Parametric Mapping software (SPM), operating under Matlab 2022b. Preprocessing steps included bias-field correction, skull stripping, and tissue segmentation into gray matter (GM), white matter, and cerebrospinal fluid, using an adaptive maximum a posteriori algorithm. GM probability maps were nonlinearly normalized to MNI152NLin2009cAsym space using CAT12’s high-dimensional geodesic shooting registration framework, modulated by the Jacobian determinant, and smoothed with an isotropic Gaussian kernel of 8 mm full-width at half-maximum.

To identify brain areas associated with homophone performance, we conducted voxel-based morphometry (VBM) analyses. Nonparametric statistical inference was performed using the Statistical NonParametric Mapping (SnPM13) toolbox implemented in SPM12 and MATLAB.(Ashburner & Friston, 2000; Nichols & Holmes, 2002) Voxel wise regression models were employed to assess the association between gray matter volume and the performance of homophone scores, with individual performance scores as the primary covariate and age at examination, sex, total gray matter volume, years of education, and testing language (Mandarin or Cantonese) included as covariates. Eight models were conducted in total. Two full-cohort models examined mono-character and bi-character accuracy separately across all participants. Six additional variant-specific models examined mono-character and bi-character accuracy separately within samples comprising cognitively healthy controls and participants with svPPA, lvPPA, or nfvPPA. Guided by our a priori hypothesis, the analyses focused on positive associations between gray matter volume and homophone selection accuracy. Significance was assessed using 5,000 permutations, with a cluster-forming threshold of t = 3.09 (uncorrected p < 0.001) and cluster-level family-wise error (FWE) correction at p < 0.05.(Friston et al., 1996)

## Results

### Demographic and neurolinguistic profiles

This study included 129 participants (66 cognitively healthy controls, 23 individuals with lvPPA, 23 with nfvPPA, and 17 with svPPA). The mean age at examination differed across diagnostic groups (F(3,125)=4.50, p=0.005), with controls being younger on average (63.2±8.2 years) than participants with lvPPA (69.0±6.9 years; p=0.011). Years of education also varied across groups (H(3)=27.81, p<0.001), with controls (16.0 [4.0]) having more years of education than participants with lvPPA (12.0 [8.0]) and nfvPPA (12.0 [6.0]). MMSE scores differed markedly across groups (H(3)=78.70, p<0.001), with controls (30.0 [2.0]) outperforming lvPPA (20.0 [6.0]), nfvPPA (26.0 [10.0]), and svPPA (22.0 [6.0]) (all p<0.001). Sex/Gender distribution also varied across groups (χ²(3)=14.54, p=0.002), whereas handedness did not (χ²(6)=6.78, p=0.341) (Table 1).

In terms of neurolinguistics features, we found differences across study groups on tasks heavily reliant on semantic knowledge, specifically the Confrontation Naming Test (CNT; H(3)=86.44, p<0.001), the Semantic Word Comprehension (SWC) test (H(3)=69.33, p<0.001), and the semantic association tasks involving pictures (H(3)=70.95, p<0.001) and words (H(3)=69.13, p<0.001) (Table 1). Controls performed near ceiling, whereas individuals with svPPA exhibited reduced performance on CNT (12.5% [16.67]), SWC (44.4% [26.67]) and both picture (66.7% [20.0]) and word association tasks (66.7% [25.0]). Post hoc comparisons also revealed that svPPA group performed worse than controls (all p<0.004) and nfvPPA across all semantic measures (p ≤ 0.046) and was also worse than lvPPA on the SWC test (p = 0.046). Moreover, group differences were also evident on all three motor-speech tasks: place of articulation (H(3)=42.91, p<0.001), manner of articulation (H(3)=45.98, p<0.001), and tone of articulation (H(3)=33.04, p<0.001). Controls performed highest across tasks: place (89.5% [15.0]), manner (95.0% [8.25]), and tone (96.2% [12.25]). Both lvPPA and nfvPPA showed impairments relative to controls on repeating place (lvPPA = 66.0% [34.5], nfvPPA = 64.0% [35.3]), manner (lvPPA = 73.5% [18.0], nfvPPA = 64.5% [44.25]) and tone of articulation phrases (lvPPA = 77.0% [19.76], nfvPPA = 64.5% [34.25]) (all p < 0.001). Additionally, nfvPPA performed worse than svPPA on repeating tone of articulation phrases (p = 0.011). Sentence repetition accuracy also differed by groups (H(3)=57.18, p<0.001) with controls sognificantly outperforming all PPA variants (Control= 89.8% [7.2], lvPPA = 67.2% [28.3], nfvPPA = 74.9% [21.0], and svPPA = 78.0% [30.5]; all p ≤ 0.004). Among the PPA groups, lvPPA showed the lowest repetition accuracy, differing from svPPA before adjustment (p = 0.043) but not after Bonferroni correction (p = 0.26). Orthographic dictation accuracy differed significantly across groups (H(3)=73.71,p<0.001), with controls achieving higher accuracy (median [IQR], 91.18% [8.82]) than the svPPA (20.59% [34.56]), nfvPPA (41.18% [52.94]), and lvPPA groups (8.82% [11.76]; all adjusted p<0.001). Syntax Comprehension likewise differed across groups (H(3)=34.86, p<0.001), with controls performing better (100% [6.67]) than the svPPA (86.67% [10.00]), nfvPPA (90.00% [18.33]), and lvPPA groups (83.33% [17.50]; all adjusted p≤0.006).

### Homophone selection performance

Figure 1 depicts observed homophone selection accuracy across diagnostic groups, whereas Table 2 reports the corresponding covariate-adjusted estimated marginal means by diagnostic group and testing language. First of all, group differences emerged across all homophone scores, specifically total, mono-character, and bi-character accuracy. For the total homophone score, Kruskal-Wallis tests revealed a robust effect of diagnosis (H(3)=65.09, p<0.001), with controls performing near ceiling (100.0% [6.25]) and all PPA groups showing marked impairments (lvPPA: 70.0% [35.0]; nfvPPA: 87.5% [30.0]; svPPA: 65.0% [46.9]; all p<0.001). Similarly, mono-character homophone performance differed across study groups (H(3)=45.97, p<0.001). In particular, controls (100.0% [0.0]) performed better than lvPPA (80.0% [60.0]), nfvPPA (75.0% [50.0]), and svPPA (62.5% [50.0]; all p<0.001). A similar pattern emerged for bi-character homophones (H(3)=67.78, p<0.001), where controls showed better performance than all PPA (Control=100.0% [0.0]; lvPPA=70.0% [50.0]; nfvPPA=87.5% [20.0]; svPPA=60.0% [30.0]; all p≤0.002). Pairwise tests also revealed that svPPA performed worse than nfvPPA (p=0.043).

**Figure 1.**
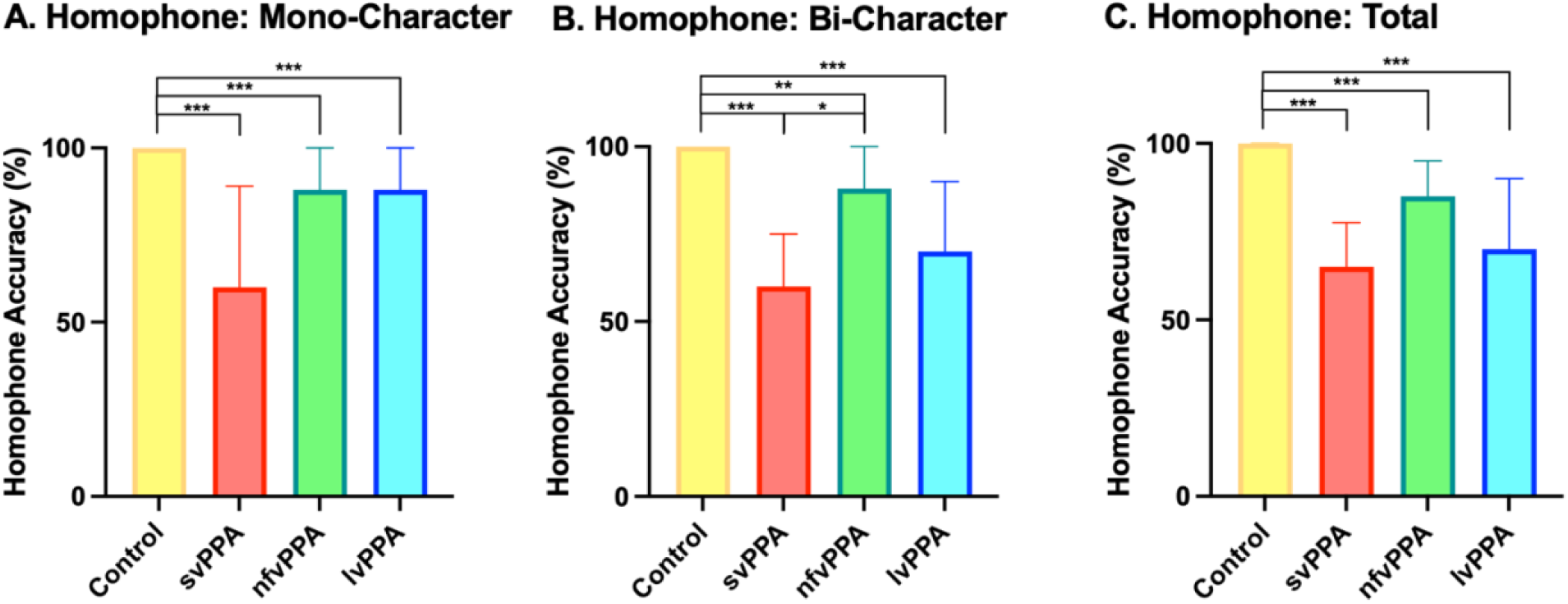
Unadjusted homophone-selection accuracy across diagnostic groups. Observed accuracy is shown for (A) mono-character, (B) bi-character, and (C) total homophone selection across cognitively healthy controls and the three PPA variants. Bars represent group means, and error bars represent [standard deviations/standard errors]. Statistical significance was evaluated using Kruskal–Wallis tests followed by Bonferroni-adjusted pairwise comparisons. Brackets denote significant pairwise differences: *p < .05, **p < .01, ***p < .001, and ****p < .0001.

**Table 2.** Covariate-adjusted estimated marginal means for homophone-selection accuracy by diagnostic group and testing language.

|  | Homophone: Mono-Character |  |  | Homophone: Bi-Character |  |  | Homophone: Total |  |  |
| --- | --- | --- | --- | --- | --- | --- | --- | --- | --- |
|  | Overall | Mandarin | Cantonese | Overall | Mandarin | Cantonese | Overall | Mandarin | Cantonese |
| Control | 98 (95-99)<sup>a,b,c</sup> | 100 (98-100)<sup>a</sup> | 86 (80-90)<sup>a,b</sup> | 98 (96-99)<sup>a,b,c</sup> | 99 (97-100)<sup>a,b,c</sup> | 96 (91-98)<sup>a</sup> | 97 (96-98)<sup>a,b,c</sup> | 99 (98-100)<sup>a,c</sup> | 91 (88-93)<sup>a</sup> |
| svPPA | 61 (48-72)<sup>a</sup> | 68 (46-100)<sup>a</sup> | 53 (43-63)<sup>a</sup> | 56 (40-70)<sup>a,d</sup> | 59 (44-72)<sup>a</sup> | 53 (28-76)<sup>a</sup> | 51 (35-66)<sup>a,d</sup> | 62 (45-77)<sup>a</sup> | 39 (18-64)<sup>a,d</sup> |
| nfvPPA | 78 (67-86)<sup>b</sup> | 87 (73-94) | 65 (52-75)<sup>b</sup> | 87 (80-92)<sup>b,d,f</sup> | 83 (72-91)<sup>b</sup> | 90 (81-95) | 82 (74-88)<sup>b,d</sup> | 84 (72-92) | 79 (70-87)<sup>d</sup> |
| lvPPA | 73 (60-83)<sup>c</sup> | 84 (70-92) | 59 (40-76) | 70 (57-80)<sup>c,f</sup> | 71 (59-80)<sup>c</sup> | 69 (46-85) | 72 (60-81)<sup>c</sup> | 76 (64-85)<sup>c</sup> | 68 (47-84) |

Superscripts denote statistically significant post hoc pairwise differences based on Šidák-adjusted p values <sup>a</sup> Significant between Control and svPPA; <sup>b</sup> Significant between Control and nfvPPA; <sup>c</sup> Significant between Control and lvPPA; <sup>d</sup> Significant between svPPA and nfvPPA; <sup>e</sup> Significant between svPPA and lvPPA; <sup>f</sup> Significant between nfvPPA and lvPPA

After accounting for age, education and testing language effect, total homophone score continued to show robust main effects of diagnosis group (Wald χ² (3)=82.34, p<0.001) and testing language (Wald χ² (1) =14.88, p<0.001), as well as an interaction between diagnosis group and testing language (Wald χ² (3) =14.92, p=0.002). Estimated marginal means (EMMs) indicated highest accuracy in controls (97%), followed by nfvPPA (82%), lvPPA (72%), and svPPA (51%). Pairwise comparison with Sidak correction confirmed that controls outperformed each PPA variant (all adjusted p≤0.001) and that svPPA scored worse than nfvPPA (adjusted p=0.001). Overall, Mandarin yielded higher accuracy than Cantonese (EMMs: 88% vs 73%, p<0.001). The language effect varied by diagnosis (interaction p=0.002) and was significant within controls (99% vs 91%, p<0.001), but not within the other groups in this model.

In the mono-character analysis, significant main effects of diagnosis group (Wald χ² (3)=38.03, p<0.001) and testing language (Wald χ² (1) =29.60, p<0.001) with a significant interaction (Wald χ² (3) =10.38, p=0.016) were noted. Overall, Mandarin outperformed Cantonese (93% [2.1] vs 67% [3.1]; p<0.001). Further pairwise comparisons showed a Mandarin advantage in controls (100% [0.2] vs 86% [2.5]; p<0.001) and svPPA (68% [10] vs 53% [5.3]; p=0.038), with non-significant trends in lvPPA (84% [5.5] vs 59% [9.6]; p=0.398) and nfvPPA (87% [5.1] vs 65% [5.9]; p=0.299). On the other hand, the bi-character homophone performance was predicted by diagnosis groups (Wald χ²=68.22, p<0.001) with no significant effect for testing language (p=0.350) or the interaction between diagnosis group and testing language (p=0.058).

Within the pooled PPA sample, homophone selection accuracy was positively correlated with semantic association: word (ρ = 0.718, n = 56), semantic association: picture (ρ = 0.660, n = 58), single word comprehension (ρ = 0.635, n = 55), confrontation naming (ρ = 0.600, n = 55), orthographic dictation (ρ = 0.550, n = 52), and syntax comprehension (ρ = 0.531, n = 48). All six associations remained statistically significant after Holm correction across the ten correlations (all Holm-adjusted p ≤ 0.010; Figure 2). The association between homophone-selection accuracy and sentence repetition was smaller and did not remain significant after correction (ρ = 0.267, n = 56, unadjusted p = 0.046, Holm-adjusted p = 0.184). Homophone-selection accuracy was not significantly associated with motor speech: place of articulation (ρ = 0.103, n = 42, unadjusted p = 0.517), motor speech: manner of articulation (ρ = −0.048, n = 42, unadjusted p = 0.763), or motor speech: tone of articulation (ρ = −0.044, n = 44, unadjusted p = 0.778); the Holm-adjusted p value was 1.000 for each association. Overall, this pattern is consistent with homophone selection drawing on semantic knowledge, lexical comprehension, orthographic processing, and comprehension of the disambiguating sentence context, while showing little association with motor-speech or articulatory performance.

**Figure 2.**
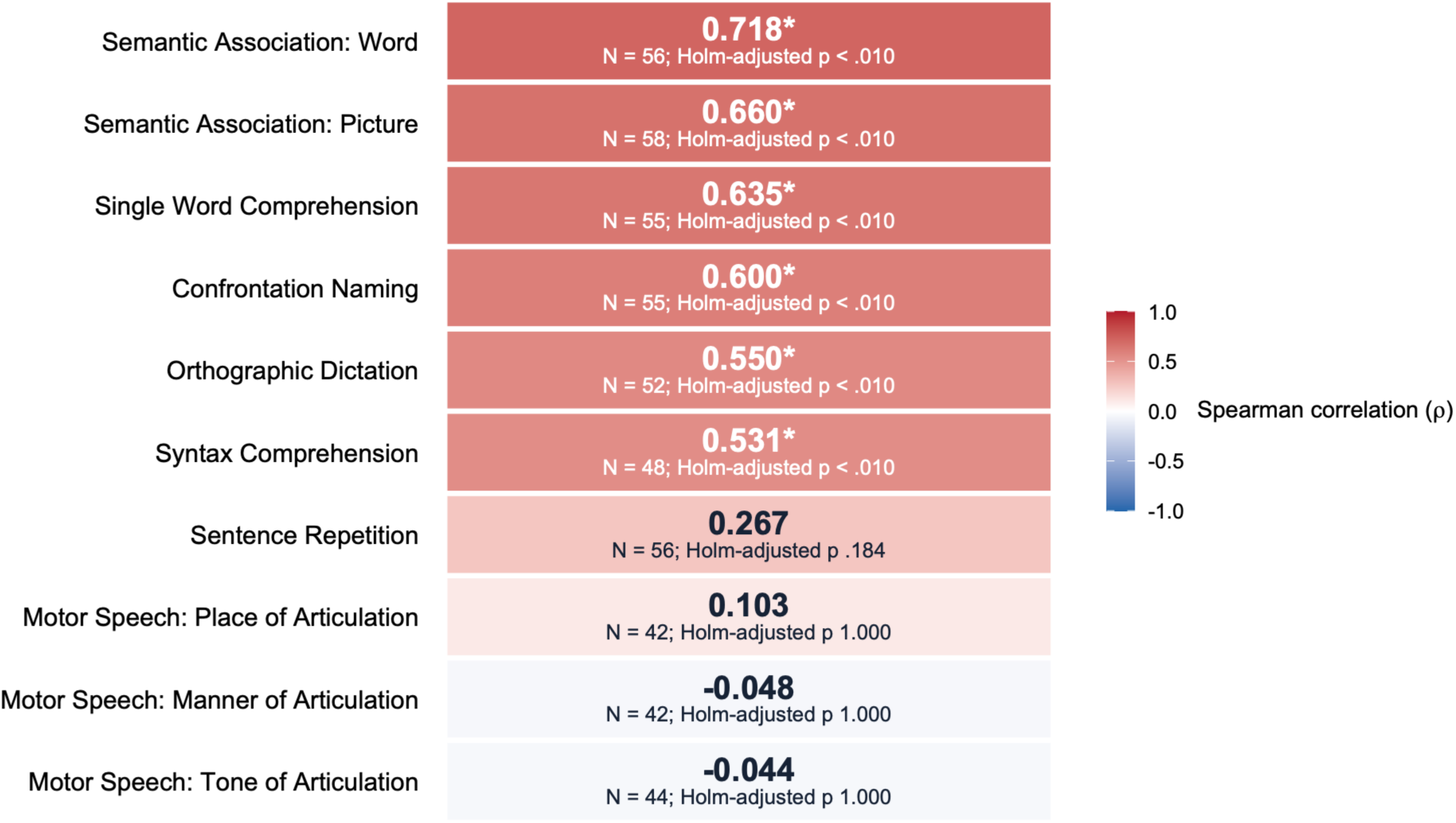
Neuropsycholinguistic correlates of homophone-selection accuracy in PPA. Heatmap showing Spearman correlations between homophone selection accuracy and neuropsycholinguistic measures in the pooled PPA sample. Sample sizes vary because correlations were calculated using pairwise available observations. Asterisks indicate associations that remained significant after Holm correction across ten comparisons.

ROC analyses were first conducted using unadjusted accuracy scores (Supplement Figure 2). For mono-character homophone accuracy, the unadjusted ROC analysis yielded an AUC of 0.809 (95% CI: 0.739–0.878). Using Youden index, the optimal cutoff was identified at 95% accuracy on the mono-character homophone task, corresponding to a sensitivity of 0.762 and a specificity of 0.788. Bi-character homophone accuracy showed slightly stronger discrimination, with an unadjusted AUC of 0.859 (95% CI: 0.797–0.920); the optimal threshold was also at 95% accuracy, producing a sensitivity of 0.790 and specificity of 0.833. We next evaluated separate multivariable models incorporating mono-character or bi-character accuracy together with age, sex, education, and testing language (Figure 3B). Both models demonstrated strong discrimination between PPA and controls (mono-character: AUC = 0.891, 95% CI [0.833, 0.949]; bi-character: AUC = 0.916, 95% CI [0.867, 0.965]). Although the AUC was numerically higher for the bi-character model, the difference was not statistically significant (ΔAUC = 0.025, 95% CI [-0.033, 0.083], z = 0.85, p = 0.396).

**Figure 3.**
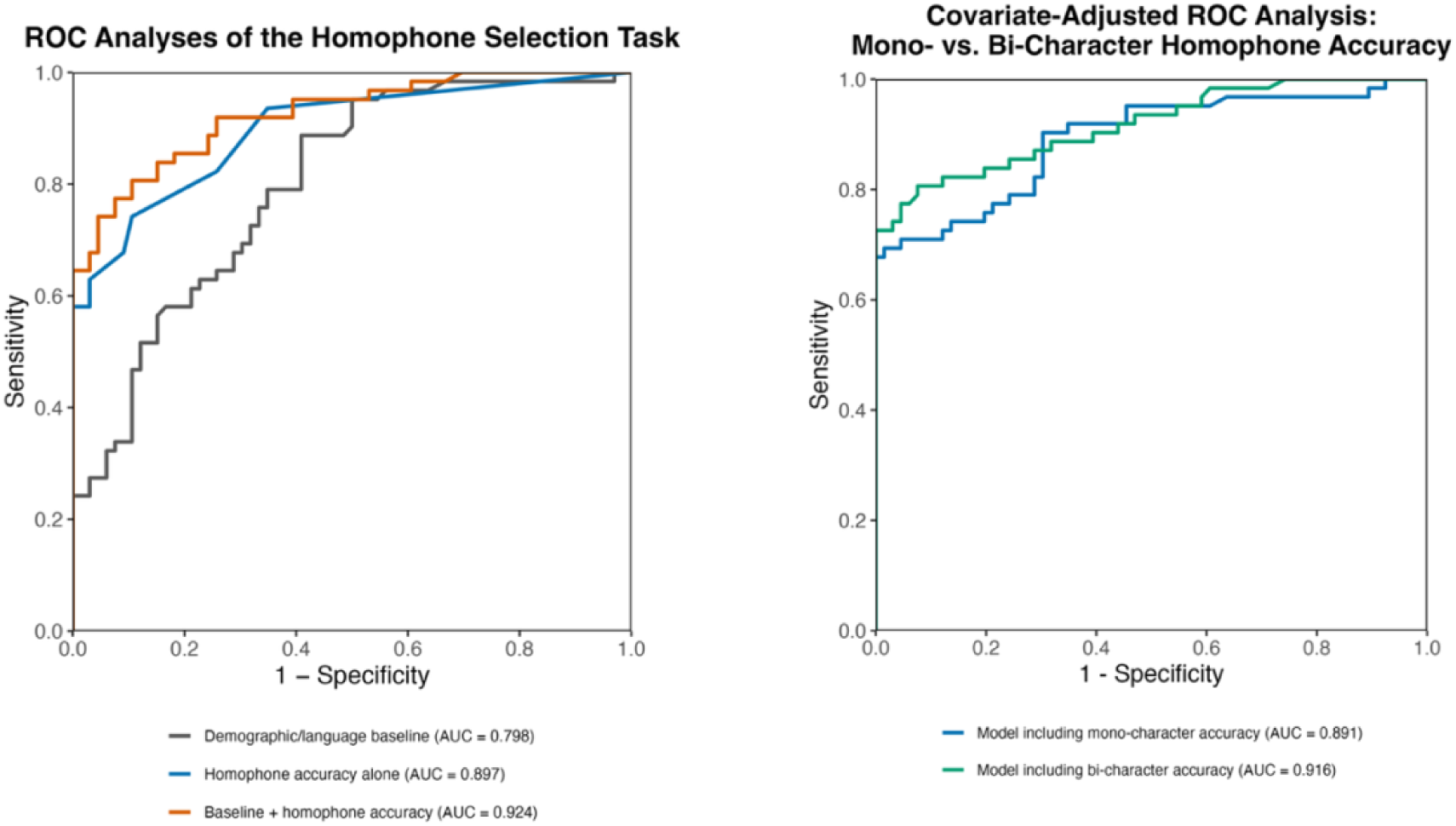
Receiver operating characteristic analyses of the homophone-selection task. (A) ROC curves compare the discrimination between participants with PPA and cognitively healthy controls provided by a baseline model containing age, sex, education, and testing language; total homophone-selection accuracy alone; and a full model combining homophone accuracy with the baseline variables. The respective AUCs were 0.798 (95% CI [0.722-0.873]), 0.897 (95% CI [0.844-.950]), and 0.924 (95% CI [0.880, 0.969]). Adding homophone accuracy to the baseline model significantly increased the AUC by 0.127 (95% CI [0.060, 0.194], paired DeLong p < .001). (B) Covariate-adjusted ROC curves compare separate multivariable models incorporating mono-character or bi-character homophone accuracy together with age, sex, education, and testing language. Both models demonstrated strong discrimination (mono-character: AUC = 0.891, 95% CI [0.833, 0.949]; bi-character: AUC = 0.916, 95% CI [0.867, 0.965]), with no significant difference between their AUCs (ΔAUC = 0.025, 95% CI [-0.033, 0.083], paired DeLong p = 0.396).

To determine whether total homophone selection accuracy provided incremental discrimination beyond demographic characteristics, we compared a baseline model containing age, sex, education, and testing language with a full model additionally containing total homophone accuracy (Figure 3A). Total homophone accuracy alone demonstrated strong discrimination (AUC = 0.897, 95% CI [0.844, 0.950]). The baseline demographic model achieved an AUC of 0.798 (95% CI [0.722, 0.873]), which increased to 0.924 (95% CI [0.880, 0.969]) after homophone accuracy was added. This improvement was statistically significant (ΔAUC = 0.127, 95% CI [0.060, 0.194], z = 3.71, p < 0.001), as was the improvement in overall model fit (likelihood-ratio Δχ²(1) = 55.76, p < 0.001; n = 128). To contextualize its discriminative performance across measurements, total homophone-selection accuracy was compared with ten neuropsycholinguistic measures. Across paired samples, total homophone selection accuracy demonstrated strong PPA-versus-control discrimination (AUC = 0.882-0.914), comparable to that of the ten neuropsycholinguistic measures examined (AUC = 0.798-0.968). Although nominal differences were observed relative to Syntax Comprehension and Orthographic Dictation, none of the paired AUC differences remained significant after Holm correction (all adjusted p ≥ 0.360; Supplementary Table 4), indicating that the discriminative performance of the homophone-selection test was comparable, but not superior, to the other measures.

### Brain Behavioral Correlation

Figure 4 and Supplementary Table 5 illustrate the gray matter volume correlates of homophone selection performance for mono-character and bi-character items, shown for the full cohort as well as for each PPA variant separately. In the all-participant analyses, we found that lower mono-character accuracy was associated with lower gray matter volume predominantly in the left inferior temporal, hippocampal, and fusiform gyri, extending into the insula and middle temporal gyrus. Similarly, bi-character accuracy showed positive correlations in these regions but with a more extensive distribution, encompassing additional clusters in the left inferior frontal and anterior temporal gyri.

**Figure 4.**
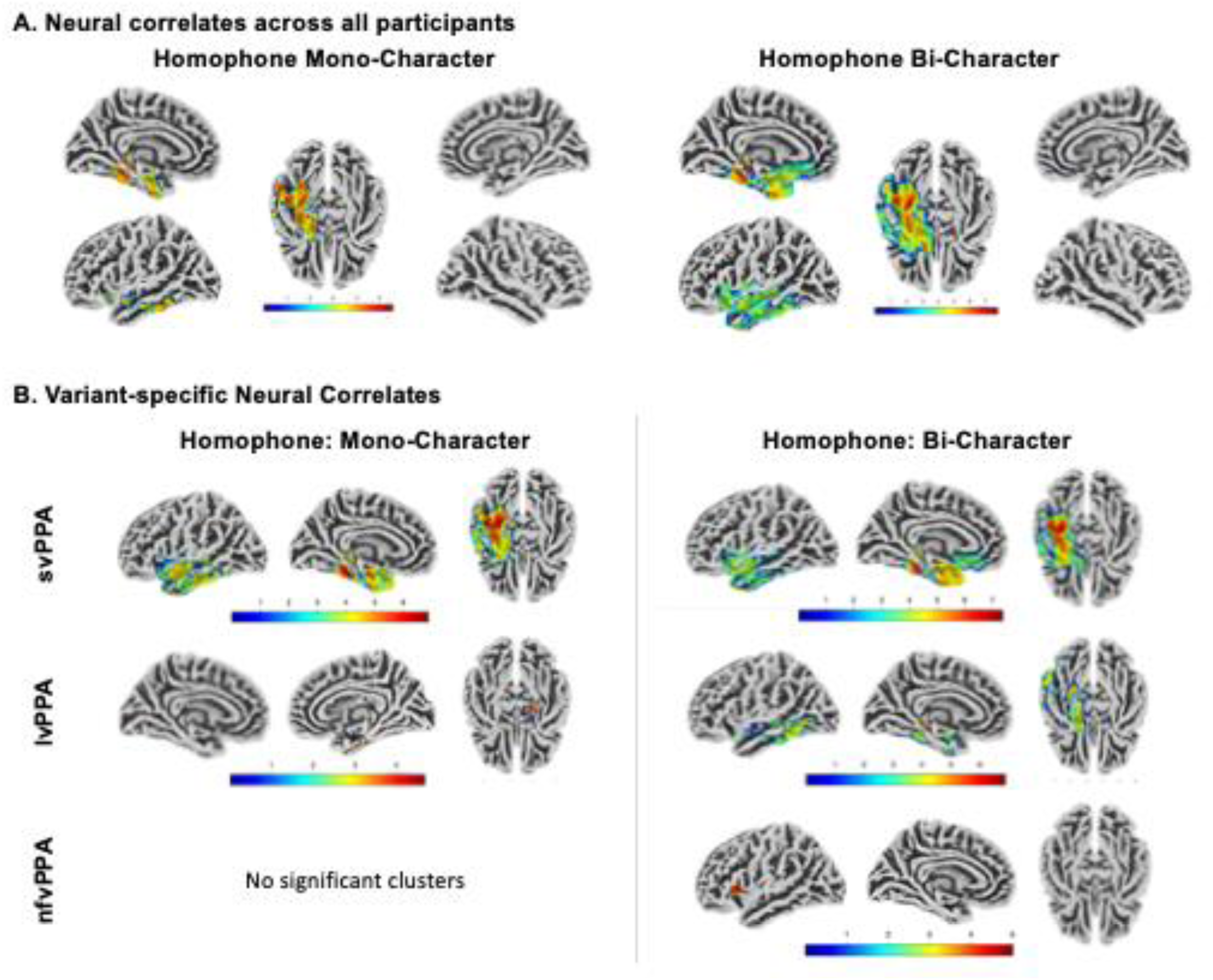
Neural Correlates of Homophone Processing in Chinese Speakers With PPA. **A. Group-level neural correlates across all participants (controls + all PPA variants).** Top Panel shows gray matter regions with significant positive correlation with homophone selection scores (Left: Mono-Character, Right: Bi-Character). Statistical non-parametric mapping (SnPM13) regression models were adopted and included homophone accuracy as the predictor of interest, controlling for age, sex, years of education,testing languages, and total gray matter volume. Statistical maps were derived from volumetric analyses and projected onto cortical surfaces for visualization. Maps are thresholded at t ≥ 3.09 (uncorrected p < 0.001) with cluster-level FWE correction at p < 0.05. **B. Variant-specific neural correlates of homophone performance.** Bottom panels display voxel-wise associations between gray matter volume and homophone performance separately for svPPA, lvPPA, and nfvPPA, for both mono-character (left column) and bi-character (right column) conditions. Across both homophone conditions, svPPA demonstrated significant associations in the left anterior temporal, inferior temporal, and fusiform regions. In contrast, lvPPA showed bilateral hippocampal and left occipital involvement for mono-character performance, whereas bi-character performance was associated with the left posterior middle temporal and inferior temporal regions. For nfvPPA, positive correlations were observed in the left insula for bi-character scores; however, no clusters survived correction for the mono-character condition. Color bars reflect variant-specific t-value ranges. All variant-specific models were thresholded at t ≥ 3.09 (uncorrected p < 0.001) with cluster-level FWE correction at p < 0.05, except for the lvPPA mono-character analysis, which is displayed at t ≥ 3.09 (uncorrected p < 0.001) with a cluster-extent threshold of k > 200 (uncorrected).

In the variant-specific analyses, at least partially distinct neuroanatomical patterns emerged for the three PPA variants. For both mono-character and bi-character homophone selection, svPPA showed significant positive correlation with gray matter volume over left inferior temporal and fusiform regions, extending into the anterior temporal gyrus. In lvPPA, higher mono-character homophone accuracy was correlated with greater gray-matter volume in bilateral medial temporal regions, including bilateral hippocampus, right parahippocampal and right fusiform cortices. For bi-character homophones, lvPPA demonstrated more widespread correlation involving the bilateral hippocampus, left middle and inferior temporal, left fusiform, left visual association cortices. In contrast, nfvPPA did not show significant associations for mono-character accuracy, but bi-character performance correlated with gray-matter volume in a left frontoinsula regions, encompasing the insula and inferior frontal gyrus.

## Discussion

To the best of our knowledge, this is the first study that investigated how homophone processing is affected in different PPA variants, which is an essential linguistic component in Chinese and other homophone-rich languages. Our findings reveal that homophone selection ability is impaired across all PPA variants, with the most pronounced deficits observed in patients with svPPA and lvPPA. The ROC analyses demonstrated strong PPA-versus-control discrimination and showed that homophone-selection accuracy contributed information beyond demographic characteristics. Exploratory correlations further indicated that homophone selection accuracy was associated with semantic, lexical-comprehension, orthographic, and syntactic abilities, but not with motor-speech performance, consistent with the task’s reliance on orthographic-semantic access and sentence-context integration. Furthermore, brain-behavior analyses indicated the involvement of the left anterior and inferior temporal regions in homophone processing, with variant-specific differences in neural correlates contributing to this deficit. This study broadens the linguistic scope of PPA research beyond English and provides critical insight into how PPA disrupts language functions that are highly dependent on homophone processing.

In our homophone-selection task, participants used sentence context to select among written alternatives sharing the same pronunciation but differing in orthography and meaning. Prior research on homophone cognitive processing in cognitively healthy individuals showed involvement of multiple cognitive abilities, including contextual interpretation(Li & Yip, 1998; Yip & Zhai, 2018), phonological(Bao et al., 2024; Carmo-Blanco et al., 2019; Carmo-Blanco et al., 2021) and orthographic processing(Chernova, 2022), and executive control(Kadem et al., 2020; Tiali et al., 2021) across both spoken and written modalities. While assessments of homophone processing in individuals with aphasia are limited, one case report published in 1979 has documented impaired homophone performance in an English-speaking patient with progressive aphasia characterized by marked semantic deterioration, likely reflecting a case of svPPA.(Schwartz et al., 1979) In our study, we found substantial deterioration of homophone selection function across all three PPA variants, with the most pronounced impairments in svPPA, followed by lvPPA and, to a lesser extent, nfvPPA. This finding supports our speculation that accurate homophone selection relies on the coordinated engagement of multiple cognitive processes. Specifically, semantic knowledge is required for contextual interpretation, a cognitive process profoundly affected in svPPA.(Sedivy et al., 1999; Tee & Gorno-Tempini, 2019) Lexical retrieval and orthographic long-term memory support access to potential homophonic alternatives, functions that are affected in lvPPA.(Gorno-Tempini et al., 2011; Tee, Kwan-Chen, et al., 2022) Finally, executive control supports accurate semantic judgment and the suppression of competing responses, functions commonly affected in nfvPPA.(Coemans et al., 2022; Gajardo-Vidal et al., 2023)

Neuroanatomically, homophone selection accuracy was associated with gray matter volume in the left anterior and inferior temporal regions and left fusiform gyrus for both mono- and bi-character items. These areas are critical hubs supporting semantic knowledge and orthographic long-term memory, consistent with the cognitive demands for homophone processing.(Binney et al., 2010; Visser et al., 2010) In addition, distinct anatomical patterns emerged across the variant-specific model, possibly reflecting differential contributions of the cognitive mechanisms supporting homophone selection across variants. For both mono- and bi-character tasks, svPPA showed neural correlates largely overlapping with those observed in the overall cohort, including the fusiform and ventral temporal regions, but with more extensive involvement of the left anterior temporal cortex. This heightened anterior temporal involvement is consistent with the greater semantic degradation characteristic of svPPA and more predominantly contributes to its homophone impairment.(Gorno-Tempini et al., 2011; Tee & Gorno-Tempini, 2019) For nfvPPA, bi-character homophone performance correlated with gray matter volume in the left insula and inferior frontal gyrus, regions involved in executive control and semantic judgment.(Ardila et al., 2014; Jackson, 2020; Whitney et al., 2011; Zhang et al., 2021) This pattern suggests that homophone impairments in nfvPPA may stem, at least in part, from difficulties with semantic control. In lvPPA, both mono- and bi-character homophone performance were associated with bilateral hippocampal regions, while bi-character performance showed additional involvement of the left fusiform gyrus and visual association cortices. The hippocampus plays a crucial role in lexical retrieval, and likely contributing to the observed homophone impairment in lvPPA.(Ludowig et al., 2008; Whitney et al., 2009) Notably, fusiform and ventral temporal involvement emerged in both the svPPA- and lvPPA-specific analyses, consistent with a shared contribution of these regions to homophone processing.

Compound words constitute approximately 70-80% of the modern Chinese lexicons.(Xing, 2006; Xue et al., 2023) For example, snowman (雪人) is a compound word formed by two meaningful morphemes (“snow雪” and “man人”). In Chinese languages, most bi-character homophones are compound words composed of two morphemes, whereas mono-character homophones are single characters that carry independent semantic meaning (e.g., snow or man) and also frequently serve as components within larger compound words.(Wang et al., 2024) Bi-character compounds help disambiguate meaning in languages with abundance homophones, as the combination of two morphemes narrows down possible homophone alternatives.(X. Chen et al., 2016; Wang et al., 2024) While homophone density is lower in bi-character compounds than mono-character homophones, patients with lvPPA in our cohort exhibited greater impairment on bi-character, accompanied by more extensive involvement of the left fusiform gyrus and visual association cortices. Given these regions are associated with visual word recognition and semantic integration, we speculate that this pattern reflects the heightened demands of orthographic analysis and semantic processing required to parse and interpret bi-character compound word forms.(Guo et al., 2022; Liu et al., 2022; Wu et al., 2012)

Notably, despite Mandarin and Cantonese sharing the same traditional Chinese written script, a language effect emerged for mono-character homophone selection, with Mandarin speakers outperforming Cantonese speakers. This dissociation may reflect the diglossic nature of Cantonese and the resulting divergence between colloquial speech and standard written Chinese, whereby spoken forms not always have direct orthographic counterparts, producing relatively low speech-writing correspondence.(Bauer, 2018; Li, 2017) This phenomenon is rarely observed in Mandarin.(Li, 2017) Consequently, accurate mono-character homophone selection in Cantonese may require greater demands on orthographic and homophone processing. In contrast, bi-character homophones showed no language effect. We contemplate that bi-character compound words present a more limited set of plausible homophone alternatives and therefore impose lower demand in homophone processing across both Cantonese and Mandarin.

Clinically, the homophone selection accuracy alone demonstrated strong discrimination between PPA and cognitively healthy controls (AUC = 0.897). More importantly, adding homophone accuracy to a baseline model containing age, sex, education, and testing language significantly increased the AUC from 0.798 to 0.924, indicating that task performance contributed discriminative information beyond demographic and language characteristics. The multivariable models incorporating mono-character and bi-character accuracy likewise demonstrated strong discrimination, with no significant difference between their AUCs. Across paired comparisons, total homophone accuracy performed comparably to the ten conventional neuropsycholinguistic measures examined, although it was not significantly superior to any measure after correction for multiple comparisons. Its clinical contribution may therefore lie not in replacing established assessments, but in complementing them by capturing impairment in a linguistically salient feature of Chinese that has rarely been characterized in PPA. Its value lies in broadening assessment to encompass language-specific processes that may be overlooked by instruments developed primarily for Indo-European languages.

Mechanistically, the behavioral and neuroanatomical findings provide converging evidence that homophone selection depends on the integration of semantic knowledge, orthographic representation, and sentence comprehension. Accuracy was associated with semantic, orthographic, and syntactic abilities, but not with motor-speech performance, while the neuroanatomical analyses implicated left temporal and fusiform regions supporting semantic and orthographic processing. The distinct patterns observed across PPA variants further suggest that impaired homophone selection may arise through different pathways of network degeneration. Although these associations do not isolate the contribution of each component process, they advance a more linguistically informed account of how neurodegeneration processes in PPA disrupts homophone selection in Chinese speakers.

While this study offers novel insights into homophone processing in Chinese-speaking individuals with PPA, several limitations should be acknowledged. Prior research has shown cross-linguistic differences in lexical decision and word learning performance for homophone versus non-homophone words and this is speculated to arise from variation in the prevalence and density of homophones across languages.(Lu & Morgan, 2020) While our study showcased substantial homophone processing impairments in Chinese PPA, further investigation is needed to examine whether comparable deficits would emerge in languages with fewer homophones and therefore lower demands on lexical disambiguation. Furthermore, although the sentence contexts were brief, predominantly composed of high-frequency words, and semantically constrained, performance required both sentence comprehension and access to orthographic-semantic representations. The present task is not positioned to distinguish impairments in these processes or their integration, and the correlation analyses do not establish their independent contributions. In addition, the identified neural correlates were necessarily constrained by the distribution of atrophy within each PPA variant and may therefore underrepresent regions that support homophone selection but remain relatively preserved. Although the spatial distributions appeared distinct across PPA variants and between mono- and bi-character homophones, suggesting potentially different anatomical contributions, these variant-specific differences were not directly tested and require confirmation through larger samples. Moreover, all study participants were native speakers of at least one Chinese languages, with most acquiring Mandarin or Cantonese natively or early in life. All but one were bilingual or multilingual. Variation in language profiles was not fully modeled and may have influenced homophone selection performance and these effects warrant investigation in larger samples. Notably, while we adopted separate but structurally parallel Mandarin and Cantonese homophone task versions and testing language was explicitly modeled, pooling the two language groups assumes comparability of the underlying homophone-selection construct. Differences in phonology, the relationship between colloquial speech and standard written Chinese may not have been fully captured. Larger language-specific samples are needed to validate measurement comparability and characterize variant effects separately in Mandarin and Cantonese. Finally, our task included only a small set of mono- and bi-character homophone items. Future studies incorporating larger and more diverse homophone sets that vary in lexical properties, semantic transparency, and orthographic complexity will be valuable for further refining our understanding of homophone processing.

## Conclusion

In summary, Chinese PPA patients exhibit significant homophone impairments, and homophone selection proves to be a an effective tool for distinguishing PPA from cognitively healthy individuals. By examining a linguistic feature that is central to many East and Southeast Asian languages, this study provides an additional clinically informative marker for PPA and offers a cross-linguistic perspective on how the disease disrupts language networks and reveals patterns of vulnerability not readily observable in Indo-European languages. Beyond its clinical value, this work illustrates how typological differences shape the neurocognitive architecture of language and underscores the importance of linguistically informed cognitive research and diagnostic approaches.

## Acknowledgements

We would like to thank the participants and their families for their generous contributions to this work. We are also grateful to Jana Klein, Xin Wen, Anna Yao, Xinge Zhao, Joanne Wu and Yun-Ting Wang at UCSF; Clara Cheng, Tammy Lau, and Kerwin Cheung in Hong Kong; as well as Yu-Ruei Lin, Yun-Ching Lo and Yu-Chung Su in Taiwan for their invaluable assistance with data collection and entry.

## Data availability

Anonymized data may be made available upon request to the corresponding author, contingent upon institutional approval and execution of a Material Transfer Agreement. Data are not publicly accessible in accordance with institutional ethical protocols governing participant confidentiality.

