## Supplementary Tables and Figures for "When Words Sound Alike: Decoding Homophone Processing in Chinese Primary Progressive Aphasia"

**Supplementary Figure 1**: Variant-specific cortical-thickness patterns in Chinese-speaking PPA.

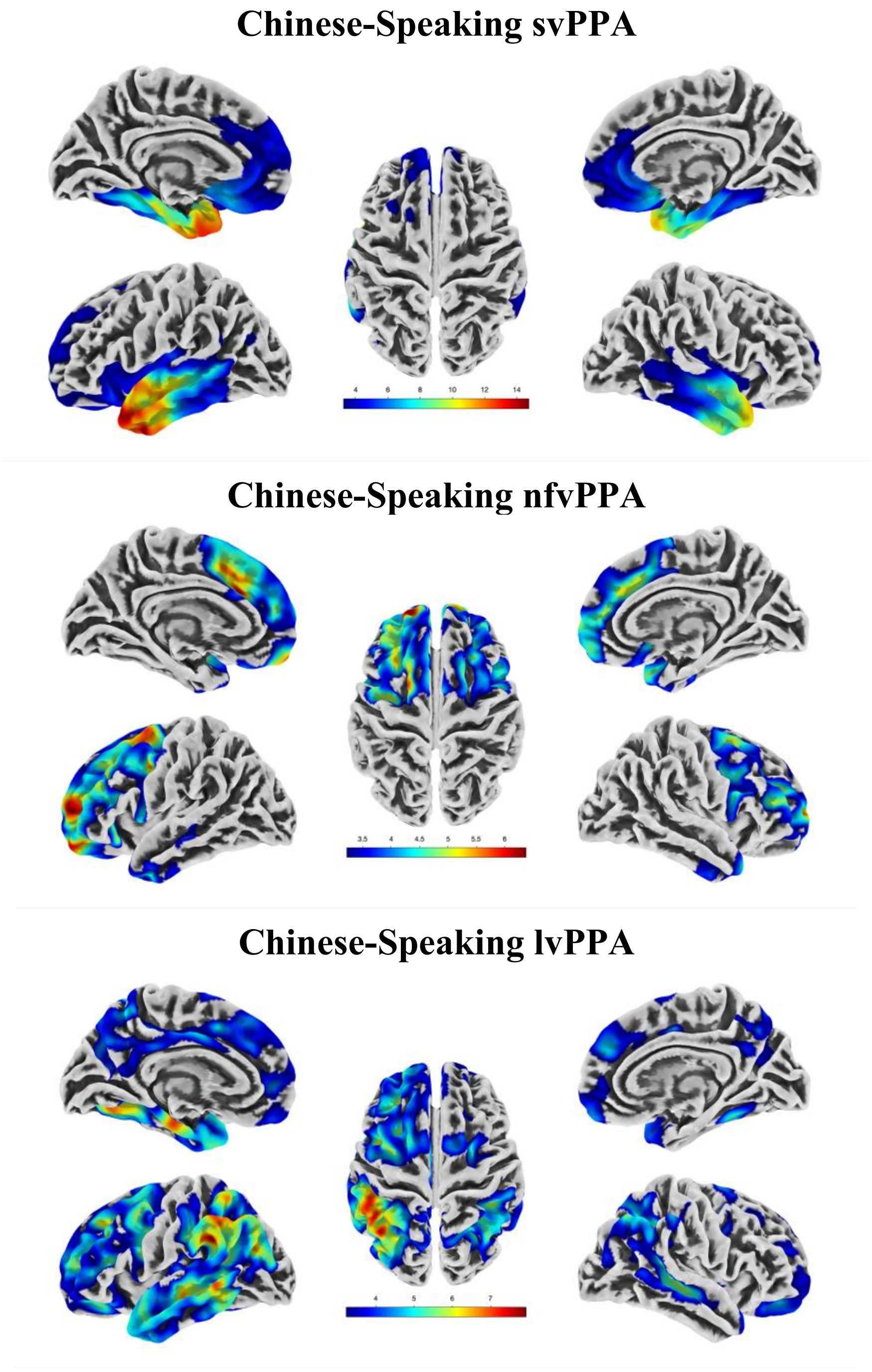


**Supplementary. Figure 1**: Variant-specific cortical-thickness patterns in Chinese-speaking PPA. Cortical-thickness differences between cognitively healthy controls and participants with svPPA, nfvPPA, and lvPPA in the whole CLAP cohort are shown after adjustment for age and sex. svPPA demonstrated predominant anterior temporal thinning, nfvPPA showed frontal-insular and perisylvian thinning, and lvPPA exhibited temporoparietal thinning.

**Supplementary Table 1. Example of High Homophone Density in Mandarin: 72 Distinct Characters Sharing the Pronunciation /Yì/.**

| Character | Pinyin | English meaning | Character | Pinyin | English meaning | Character | Pinyin | English meaning |
|---|---|---|---|---|---|---|---|---|
| 意 | yì | meaning; intention | 翊 | yì | assist; support | 瘞 | yì | skin blemish |
| 义 / 義 | yì | righteousness; justice | 佚 | yì | lost; idle | 饐 | yì | spoiled food |
| 亿 / 億 | yì | hundred million | 屹 | yì | towering; upright | 痍 | yì / yí | wound; injury |
| 议 / 議 | yì | discuss; deliberate | 役 | yì | service; labor | 謚 | yì | posthumous title |
| 易 | yì | easy; change | 亦 | yì | also | 邘 | yì / yú | ancient place name |
| 异 / 異 | yì | different; unusual | 奕 | yì | grand; abundant | 匜 | yì / yí | ladle; basin |
| 译 / 譯 | yì | translate | 弈 | yì | play chess (Go) | 眙 | yì / yí | stare; gaze |
| 谊 | yì | friendship; goodwill | 怡 | yí | joyful; harmonious | 嗣 | yì / sì | heir; inherit |
| 翌 | yì | next (day) | 翳 | yì | shade; screen | 繶 | yì / yù | bind; tie |
| 益 | yì | benefit; increase | 熤 | yì / yù | glowing flame | 螠 | yì | beetle |
| 邑 | yì | city; town | 熼 | yì | blazing fire | 螘 | yì / yǐ | ant |
| 翼 | yì | wing; assist | 峄 / 嶧 | yì | Mt. Yi (place name) | 蟻 | yì / yǐ | ant |
| 毅 | yì | resolute; firm | 殔 | yì | punishment (rare) | 檍 | yì | tree name |
| 熠 | yì | glittering; shining | 瘗 / 瘞 | yì | bury | 薏 | yì | Job’s tears (coix seed) |
| 诣 / 詣 | yì | go to; attain | 曀 | yì | dark; overcast | 齮 | yì / yǐ | bite; gnaw |
| 裔 | yì | descendant | 嶷 | yì / yí | thoughtful; hesitant | 炨 | yì | blazing fire |
| 驿 / 驛 | yì | relay station; post | 潩 | yì | river name | | | |
| 殪 | yì | kill; put to death | 澺 | yì | river name | | | |
| 懿 | yì | virtuous; admirable | 溰 | yì | deep water | | | |
| 疫 | yì | epidemic | 藙 | yì | strong; hardy | | | |
| 羿 | yì | Yi (legendary archer) | 燡 | yì | bright flame | | | |
| 镒 / 鎰 | yì | ancient weight unit | 釔 | yì / yǐ | yttrium | | | |
| 嗌 | yì | throat; choke | 艺 / 藝 | yì | art; skill | | | |
| 燚 | yì | blazing fire | 繹 | yì | unravel; explain | | | |
| 肄 | yì | practice; study | 鷁 | yì | mythical water bird | | | |
| 曳 | yì | drag; pull | 鶍 | yì | pheasant-like bird | | | |
| 佾 | yì | ritual dancers (row) | 鷧 | yì | phoenix-like bird | | | |
| 挹 | yì | scoop; ladle | 癔 | yì | hysteria | | | |

**Supplementary Table 2. Homophone Selection Test in Mandarin and Cantonese.**

| Sentence Completion Paradigm | Choices | Pinyin | Target Homophone |
|---|---|---|---|
| **Mandarin Homphone Test** | | | |
| 汽車撞倒了____士。<br>*The car knocked down the ____.* | 騎 *ride* │ 旗 *flag* │ 奇 *strange* │ 齊 *uniform* | Qí | 騎 |
| 台北是濕冷的____市。<br>*Taipei is a damp and cold ____ city.* | 成 *become* │ 乘 *ride* │ 程 *process* │ 城 *city* | Chéng | 城 |
| 小孩喜歡有插____的書。<br>*Children like books with inserted ____.* | 話 *speech* │ 畫 *pictures* │ 化 *transform* │ 劃 *cut/draw* | Huà | 畫 |
| 這____戲機很好玩。<br>*This ____ game console is very fun.* | 由 *from* │ 遊 *play* │ 尤 *especially* │ 油 *oil* | Yóu | 遊 |
| 麥克風要接上電____。<br>*The microphone needs to be connected to a power ____.* | 員 *staff* │ 源 *source* │ 圓 *round* │ 元 *unit* | Yuán | 源 |
| 因為颱風，路口被____住。<br>*Because of the typhoon, the intersection was ____.* | 裹 *wrap* │ 睹 *see* │ 篤 *sincere* │ 堵 *block* | Dǔ | 堵 |
| 花瓣上凝結了____水。<br>*____ water condensed on the flower petals.* | 露 *dew* │ 祿 *fortune* │ 鷺 *egret* │ 漉 *drip* | Lù | 露 |
| 這鑽石首____很漂亮。<br>*This diamond ____ is very beautiful.* | 誓 *vow* │ 適 *suitable* │ 飾 *ornament* │ 釋 *explain* | Shì | 飾 |
| 運動對身體健康有____。<br>*Exercise has ____for physical health.* | 抑 *suppress* │ 液 *liquid* │ 藝 *art* │ 益 *benefit* | Yì | 益 |
| 請用____帶把紙箱封好。<br>*Please use ____ tape to seal the box.* | 郊 *suburb* │ 膠 *glue* │ 嬌 *delicate* │ 焦 *burnt* | Jiāo | 膠 |
| 這案子還有____。<br>*This case still has a ____.* | 引擎 *engine* │ 癮晴 *addiction–sun* │ 飲情 *drink–emotion* │ 隱情 *hidden truth* | Yǐn Qíng | 隱情 |
| ____是一種黃色水果。<br>*____ is a type of yellow fruit.* | 皮扒 *skin-peel* │ 琵琶 *lute* │ 枇杷 *loquat* │ 埤爬 *embankment-climb* | Pí Pá | 枇杷 |
| 飛機____香港飛往泰國。<br>*The plane flies ____ Hong Kong to Thailand.* | 精油 *essential oil* │ 經由 *via* │ 京遊 *capital tour* │ 驚游 *startled swim* | Jīng Yóu | 經由 |
| 運動會因下雨而____。<br>*The sport event was ____ due to rain.* | 中指 *middle finger* │ 忠紙 *loyal paper* │ 鐘只 *bell only* │ 終止 *terminate* | Zhōng Zhǐ | 終止 |
| 兩億不是小____。<br>*Two hundred million is not a small ____.* | 樹木 *trees* │ 術牧 *technique-herd* │ 束墓 *bundle-grave* │ 數目 *number* | Shù Mù | 數目 |
| 我軍發動了____。<br>*Our army launched an ____.* | 公事 *public affairs* │ 公式 *formula* │ 攻勢 *offensive movement* │ 恭世 *respectful era* | Gōng Shì | 攻勢 |
| ____久了腳會疼。<br>*After ____for a long time, your feet will hurt.* | 站立 *standing* │ 戰力 *combat power* │ 顫慄 *tremble* │ 佔麗 *occupy-beauty* | Zhàn Lì | 站立 |
| 性別____仍常見於職場。<br>*Gender ____ is still common in the workplace.* | 騎士 *knight* │ 歧視 *discrimination* │ 奇式 *odd style* │ 齊事 *uniform matter* | Qí Shì | 歧視 |

| | | | |
|---|---|---|---|
| ____說：「歡迎光臨」。<br>*The ____ said, "Welcome."* | 店員 *shop assistant* ｜ 電源 *power supply* ｜ 墊圓 *cushion-round* ｜ 殿元 *palace-unit* | Diàn Yuán | 店員 |
| ____愈大，風險愈高。<br>*The larger the ____, the higher the risk.* | 堵住 *block* ｜ 賭注 *stake* ｜ 睹助 *witness-help* ｜ 篤柱 *solid pillar* | Dǔ Zhù | 賭注 |
| **Cantonese Homophone Test** | | | |
| 這企業的__旨是誠信為本。<br>*This company's ____ is integrity-based.* | 棕 *brown* ｜ 宗 *principle* ｜ 中 *middle* ｜ 終 *end* | Jung1 | 宗 |
| 港姐的__色都好出眾。<br>*Miss Hong Kong contestants' ____ are outstanding.* | 知 *know* ｜ 資 *resources* ｜ 脂 *fat* ｜ 姿 *appearance* | Ji1 | 姿 |
| 犯人妨礙__法公正。<br>*The criminal obstructed ____ justice.* | 絲 *silk* ｜ 師 *teacher* ｜ 司 *judicial* ｜ 施 *apply* | Si1 | 司 |
| 娛樂圈新聞__息萬變。<br>*Entertainment news changes ____ rapidly.* | 瞬 *instant* ｜ 訊 *information* ｜ 遜 *inferior* ｜ 信 *trust* | Seun3 | 瞬 |
| 請唔好__負佢既好意。<br>*Please do not ____ her goodwill.* | 故 *cause* ｜ 辜 *betray* ｜ 孤 *lonely* ｜ 姑 *aunt* | Gu1 | 辜 |
| 這新聞莫__奇妙。<br>*This news is extremely ____.* | 明 *bright* ｜ 鳴 *cry* ｜ 名 *famous* ｜ 銘 *inscribe* | Meng2 | 名 |
| __點工人既人工好低。<br>*____ o'clock workers' wages are very low.* | 中 *middle* ｜ 宗 *clan* ｜ 鐘 *clock* ｜ 終 *end* | Jung1 | 鐘 |
| 導____會帶旅行團購物。<br>*Tour ____ lead groups shopping.* | 油 *oil* ｜ 游 *swim* ｜ 遊 *tour* ｜ 由 *from* | Yau4 | 遊 |
| 工匠用__ __磨平木頭。<br>*Craftsmen use ____ ____ to smooth wood.* | 沙子 *sand* ｜ 紗紙 *gauze paper* ｜ 砂紙 *sandpaper* ｜ 鯊子 *shark* | Sa1 Ji2 | 砂紙 |
| 車胎是__ __的。<br>*The tire is ____.* | 元型 *original type* ｜ 圓形 *round* ｜ 原形 *original form* ｜ 園型 *garden type* | Yun4 Ying4 | 圓形 |
| 警察__ __敏捷。<br>*Police officers are ____.* | 身手 *skill* ｜ 申首 *declare head* ｜ 新手 *rookie* ｜ 伸首 *extend head* | San1 Sau2 | 身手 |
| 他追求完美的__ __。<br>*He pursues a perfect ____.* | 景解 *scene-explain* ｜ 警戒 *alert* ｜ 經界 [a] *boundary* ｜ 境界 *realm* | Ging2 Gaai3 | 境界 |
| 政府推行__ __運動日。<br>*The government promotes a ____ campaign day.* | 全民 *all people* ｜ 傳聞 *rumor* ｜ 痊文 *healed text* ｜ 泉聞 *spring news* | Chyun4 Man4 | 全民 |
| 她健康而且__ __紅潤。<br>*She is healthy and has a rosy ____.* | 氣色 *complexion* ｜ 器識 *device knowledge* ｜ 憩息 *rest* ｜ 戲色 *play color* | Hei3 Sik1 | 氣色 |
| 他__ __有粒痣。<br>*He has a mole on his ____.* | 手掌 *palm* ｜ 叟掌 *old palm* ｜ 首長 *leader* ｜ 守漲 *guard-swell* | Sau2 Jeung2 | 手掌 |
| 傷口__ __要靜養。<br>*The wound needs to ____ and rest.* | 服完 *finish taking* ｜ 福元 *blessing origin* ｜ 復原 *recover* ｜ 復元 *restore* | Fuk6 Yun4 | 復原 |

[a] For each item, the syllabic structure is identical to the target word but tone varies in this choice (e.g., *經界* /ging1 gaai3/ vs. *境界* /ging2 gaai3/)； [b] *鐘點* means hourly-based (e.g., paid by the hour).；[c] *導遊* means tour guide

**Supplementary Table 3. Generalized Linear Model for Mono-character and Bi-Character Homophone Accuracy**

| | Homophone: Mono-Character | | | | Homophone: Bi-Character | | |
|---|---|---|---|---|---|---|---|
| **Effect** | **Wald χ²** | **df** | **p-value** | **Effect** | **Wald χ²** | **df** | **p-value** |
| Diagnosis | 38.03 | 3 | <0.001 | Diagnosis | 68.22 | 3 | <0.001 |
| Testing Language | 29.60 | 1 | <0.001 | Testing Language | 0.94 | 1 | 0.333 |
| Age at examination | 3.64 | 1 | 0.056 | Age at examination | 2.35 | 1 | 0.125 |
| Years of education | 4.58 | 1 | 0.032 | Years of education | 0.24 | 1 | 0.627 |
| Diagnosis x Language | 10.38 | 3 | 0.016 | Diagnosis x Language | 7.48 | 3 | 0.058 |

### Supplementary Figure 1. Unadjusted Receiver Operating Characteristic (ROC) Curves for Homophone Selection Task.

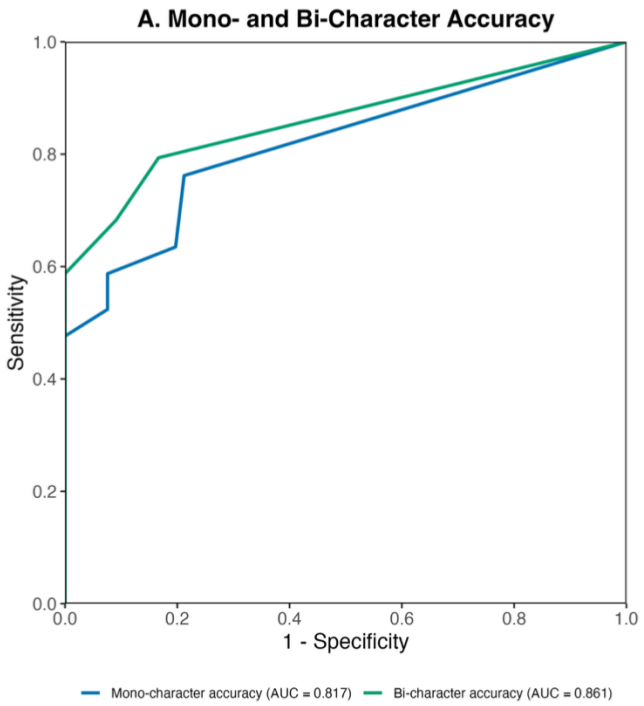

**Supplementary Table 4. Comparative discriminative performance of homophone selection test and other neuropsycholinguistic measures**

| Comparator | Paired n, PPA/control | Homophone AUC (95% CI) | Comparator AUC (95% CI) | ΔAUC (95% CI) | Raw p | Holm-adjusted p |
|---|---|---|---|---|---|---|
| Confrontation Naming | 126, 60/66 | 0.882 (.824–.940) | 0.927 (.876–.978) | −0.045 (−.113 to .023) | 0.197 | 1.000 |
| Single Word Comprehension | 117, 60/57 | 0.896 (.841–.951) | 0.890 (.835–.945) | 0.006 (−.068 to .081) | 0.872 | 1.000 |
| Semantic Association: Picture | 128, 63/65 | 0.887 (.831–.942) | 0.864 (.805–.924) | 0.022 (−.047 to .092) | 0.532 | 1.000 |
| Semantic Association: Word | 126, 61/65 | 0.883 (.826–.940) | 0.858 (.796–.921) | 0.025 (−.043 to .093) | 0.478 | 1.000 |
| Motor Speech: Place of Articulation | 84, 42/42 | 0.891 (.824–.958) | 0.893 (.827–.959) | −0.002 (−.105 to .101) | 0.970 | 1.000 |
| Motor Speech: Manner of Articulation | 84, 42/42 | 0.891 (.824–.958) | 0.901 (.828–.973) | −0.009 (−.104 to .085) | 0.846 | 1.000 |
| Motor Speech: Tone of Articulation | 86, 44/42 | 0.892 (.826–.958) | 0.798 (.708–.889) | 0.094 (−.017 to .205) | 0.099 | .792 |
| Sentence Repetition | 119, 59/60 | 0.905 (.852–.957) | 0.886 (.827–.946) | 0.018 (−.056 to .092) | 0.626 | 1.000 |
| Syntax Comprehension | 109, 49/60 | 0.914 (.863–.966) | 0.818 (.739–.897) | 0.096 (.006–.186) | 0.036 | .360 |
| Orthographic Dictation | 103, 52/51 | 0.909 (.855–.963) | 0.968 (.935–1.000) | −0.059 (−0.114 to −0.004) | 0.037 | .360 |

AUCs were calculated within the paired sample available for each comparison; consequently, the AUC for homophone-selection accuracy varies slightly across rows. ΔAUC was calculated as homophone selection AUC minus comparator AUC, such that positive values favor homophone selection. AUCs were compared using paired nonparametric tests, with Holm correction across ten comparisons. None of the paired AUC differences was statistically significant.

**Supplementary Table 5. Neuroanatomical correlates of homophone selection test.**

| Regions | Voxel-level interference | | Cluster-level interference | | Peak MNI coordinates | | |
|---|---|---|---|---|---|---|---|
| | t-value | p-value (unc) | Extent | p-value (FWE-corr) | x | y | z |
| **A. All Participants** | | | | | | | |
| **Homophone: Mono-Character** | | | | | | | |
| Left Hippocampus | 5.83 | <0.0001 | 11858 | <0.0001 | -27 | -27 | -4 |
| Left Fusiform | 5.04 | <0.0001 | | | -54 | -57 | -21 |
| Left Inferior Temporal Gyrus | 4.87 | <0.0001 | | | -48 | -38 | -22 |
| **Homophone: Bi-Character** | | | | | | | |
| Left Hippocampus | 7.78 | <0.0001 | 32414 | <0.0001 | -28 | -28 | -8 |
| Left Fusiform | 6.92 | <0.0001 | | | -46 | -39 | -21 |
| Left Fusiform | 6.86 | <0.0001 | | | -30 | -33 | -21 |
| **B. Variant-specific models** | | | | | | | |
| **Homophone: Mono-Character - svPPA** | | | | | | | |
| Left parahippocampus | 7.15 | <0.0001 | 26712 | <0.0001 | -36 | -33 | -22 |
| Left Inferior Temporal Gyrus | 6.97 | <0.0001 | | | -46 | -38 | -24 |
| Left Inferior Temporal Gyrus | 6.72 | <0.0001 | | | -50 | -30 | -20 |
| **Homophone: Mono-Character - lvPPA** | | | | | | | |
| Right Hippocampus | 4.81 | <0.0001 | 1870 | <0.0001 | 21 | -14 | -16 |
| Right Parahippocampus | 4.11 | <0.0001 | | | 22 | -14 | -39 |
| Right Fusiform | 3.35 | <0.0001 | | | 27 | -26 | -28 |
| Left Hippocampus | 4.66 | <0.0001 | 776 | 0.020 | -27 | -22 | -9 |
| **Homophone: Bi-Character- svPPA** | | | | | | | |
| Left Parahippocampus | 8.04 | <0.0001 | 25479 | <0.0001 | -38 | -32 | -22 |
| Left Fusiform | 6.96 | <0.0001 | | | -46 | -39 | -22 |
| Left Hippocampus | 6.42 | <0.0001 | | | -30 | -20 | -12 |
| **Homophone: Bi-Character - nfvPPA** | | | | | | | |
| Left Insula | 5.15 | <0.0001 | 1276 | 0.002 | -39 | 21 | -2 |
| Left Insula | 4.09 | <0.0001 | | | -26 | 22 | 3 |
| Left Pars Orbitalis | 3.85 | <0.0001 | | | -28 | 32 | -2 |
| **Homophone: Bi-Character - lvPPA** | | | | | | | |
| Left Hippocampus | 6.94 | <0.0001 | 7850 | <0.0001 | -30 | -27 | -8 |
| Left Hippocampus | 6.77 | <0.0001 | | | -33 | -20 | -12 |
| Left Hippocampus | 5.32 | <0.0001 | | | -14 | -14 | -12 |
| Left Visual Association Cortex | 4.93 | <0.0001 | 2919 | <0.001 | -52 | -66 | 2 |
| Left Middle Temporal Gyrus | 4.66 | <0.0001 | | | -63 | -48 | -6 |
| Left Visual Association Cortex | 4.58 | <0.0001 | | | -44 | -74 | 9 |
| Right Hippocampus | 4.86 | <0.0001 | 1256 | 0.002 | 36 | -12 | -16 |
| Right Hippocampus | 3.90 | <0.0001 | | | 27 | -30 | -3 |
| Right Hippocampus | 3.71 | <0.0001 | | | 39 | -30 | -2 |

The table lists representative peak voxels. FWE-corr = family-wise error corrected; MNI = Montreal Neurological Institute space; unc = uncorrected
